# Supervised self-collected SARS-CoV-2 testing in indoor summer camps to inform school reopening

**DOI:** 10.1101/2020.10.21.20214338

**Authors:** Peter Cooch, Annalisa Watson, Apryl Olarte, Emily Crawford, CLIAhub Consortium, Joe DeRisi, Bryan Greenhouse, Jill Hakim, Keirstinne Turcios, Lee Atkinson-McEvoy, Raphael Hirsch, Roberta L. Keller, Theodore Ruel, Auritte Cohen-Ross, Araceli Leon, Naomi Bardach

**Affiliations:** Department of Pediatrics, University of California, San Francisco, CA; Philip R. Lee Institute for Health Policy Studies, University of California, San Francisco, CA; Department of Microbiology and Immunology, University of California, San Francisco, CA; Department of Biochemistry and Biophysics, University of California, San Francisco, CA; Chan Zuckerberg Biohub, San Francisco, CA; Department of Medicine, University of California, San Francisco; Celsius and Beyond Science Camps, San Francisco, CA; Buena Vista Child Care, San Francisco, CA

**Author notes:** Address correspondence to: Peter B. Cooch, MD, University of California, San Francisco Department of Pediatrics, 550 16^th^ St, 4^th^ Floor, San Francisco, CA, 94143.

## Abstract

**Background and Objectives:** Testing strategies for severe acute respiratory syndrome coronavirus 2 (SARS-CoV-2) infection in school settings are needed to assess the efficacy of infection mitigation strategies and inform school reopening policies. We hypothesized that supervised serial self-collected non-nasopharyngeal testing in summer camp settings would be acceptable and feasible.

**Methods:** We performed a cohort study at two urban day camps for kindergarten-8^th^ graders in June and July 2020. Eligible participants were campers, up to two adult household contacts, and camp staff. We assessed participation rates for providing, at two time points, supervised, self-collected anterior nares samples for reverse transcription polymerase chain reaction (RT-PCR) and saliva samples for antibody testing. We qualitatively assessed testing feasibility and adherence to stated camp infection mitigation strategies.

**Results:** 76% (186/246) of eligible participants consented. The cohort completing both rounds of testing (n=163) comprised 67 campers, 76 household contacts, and 20 staff. Among those present, 100% of campers and staff completed test collection at both time points. Testing was feasible to implement, including staff participation supervising camper test collection. No virus was detected by RT-PCR; seven participants had antibodies. Observed adherence to stated camp mitigation policies for masking, physical distancing, and stable cohorting was generally high.

**Conclusions:** Supervised, self-collected serial anterior nasal and saliva-based SARS-CoV-2 testing was acceptable, with successful repeated participation by children ages 5-14. This strategy for testing and the observed infection mitigation practices comprise potential core components for safe school reopening.

## Background

All 50 U.S. states implemented school closures in response to the coronavirus disease 2019 (COVID-19) pandemic. While COVID-19 generally causes mild disease in children and adolescents, school closures are known to adversely impact children and families economically, educationally, and psychologically, while worsening race, gender, and economic disparities.^1–5^ Hence, school reopening is a priority for the well-being of children and adolescents. However, it must be done safely for the sake of teachers, staff, and student families. Testing for severe acute respiratory syndrome coronavirus 2 (SARS-CoV-2), the virus that causes COVID-19, paired with infection mitigation strategies (e.g., masking, physical distancing, stable cohorts, and hand hygiene), comprise a comprehensive strategy for safe school reopening.^6^ Symptom-based screening, and contact tracing and investigation will likely require frequent testing in children and staff.^6^ Asymptomatic surveillance, an even broader testing strategy, requires additional testing. For example, New York City is proposing monthly surveillance testing for 10% of public school students and staff.^7,8^

While school reopening will entail increased testing needs, it is unclear what testing approaches will be acceptable and feasible. Nasopharyngeal (NP) sampling, the primary SARS-CoV-2 test collection modality to date, is uncomfortable, with risk of refusal by both adults and children, requires medical provider collection, confers risk of transmission, and requires extensive personal protective equipment (PPE) including N95 mask, gown, face shield, and gloves, many of which have faced national shortages.^9,10^

Alternative sampling options include anterior nares or oral fluid (hereafter “saliva”) collection. Anterior nares collection is more comfortable than NP sampling,^11^ can be self-performed, is approved by the Centers for Disease Control and Prevention (CDC) for SARS-CoV-2 testing,^12^ with comparable sensitivity and specificity to provider-performed NP sampling,^9^ and reduces need for PPE.^9,10,13^ Non-invasive antibody testing using saliva is commonly used in HIV screening,^14^ and is being studied for SARS-CoV-2. A school-based, non-NP testing strategy for SARS-CoV-2 has not been previously published.

Additionally, the necessary extent of infection mitigation strategies in the school setting is not known, and there is debate to what degree students are able to adhere to restrictive policies, such as masking or physical distancing.

To address these questions, we evaluated serial non-NP-specimen SARS-CoV-2 testing in two summer camps. We hypothesized that supervised self-collection of anterior nares and saliva samples for the purpose of SARS-CoV-2 surveillance would be acceptable and feasible for kindergarten through 8^th^ grade children, their household contacts, and camp staff, and that camp staff could assist with collection supervision. We also observed infection mitigation practices and estimated the incidence of SARS-CoV-2 infection using reverse transcription polymerase chain reaction (RT-PCR) testing (anterior nares specimen) and antibody testing (saliva specimen) collected at the beginning and end of camp sessions.

## Methods

### Study Design and Setting

We performed a prospective cohort study at two indoor day camps within San Francisco, California: a general camp and a science camp. The general camp was 5 weeks long, was affiliated with a parochial school, enrolled kindergarten through 5^th^ grade campers, and was located in a zip code with higher COVID-19 prevalence compared to other parts of San Francisco. The science camp was 3 weeks long, enrolled 2^nd^ through 8^th^ grade campers, and was located in a zip code with lower COVID-19 prevalence. Camps were in session June-July 2020.

### Participants

Eligible participants were: all children attending the summer camps (“campers”); up to two adults from each camper household (“household contacts”); and camp staff working the entire session (“staff”). Study participant-facing materials encouraged inclusion of household contacts who worked outside the home during the pandemic. We excluded participants from the final cohort who were not present for both test collection sessions.

### Recruitment and enrollment

Camp directors sent a recruitment email to parents and staff, including a link to the consent form and baseline survey and a video of a child self-collecting an anterior nasal swab (Supplement A). The study team obtained electronic or paper consent in English or Spanish. There was no compensation for participation. We informed participants that they would receive their results from RT-PCR but not antibody testing.

### Co-variates

The baseline survey collected participant characteristics. Adult participants (household contacts and staff) completed the survey on their behalf and on behalf of participating campers. For all participants, we gathered age, gender, race, ethnicity, home zip code, and cohabitation with confirmed or suspected COVID-19 cases. From campers and camp staff we elicited a list of potential COVID-19 symptoms within 14 days prior to testing. Household contacts reported their highest level of education, whether they continued to work outside the home during the pandemic (“frontline worker”), and if so, what category of work. From state databases, we obtained participant household zip code-level cumulative incidence of COVID-19 at camp start (June 19-26, 2020).^15^ For households in a county that did not report zip code incidence (n=8), we used the households’ city cumulative incidence.

### Specimen collection, handling, and testing assays

We collected samples at two time points: within the first three and last two days of the camp session. Collection took place in an outdoor setting or in large classrooms with no more than three participants at a time, following CDC recommendations for distanced collection and specimen handling.^13^ Maintaining 6 feet of distance, the research team instructed participants to self-collect an anterior nares swab and a saliva swab (Supplements B and C).

We offered camp staff the opportunity to assist with supervision of camper specimen collection. On-site training involved observing and participating in self-collection. Staff demonstrated self-collection to campers and then helped observe and coach them. No staff independently led camper specimen collection without research team supervision, to assure standardized and adequate specimen collection.

We assessed active SARS-CoV-2 infection by RT-PCR and prior exposure by antibody testing. RT-PCR of viral N and E genes and human RNAse P gene was performed on the anterior nares samples at the Chan Zuckerberg Biohub at the University of California, San Francisco (UCSF). Saliva samples were tested for IgG at UCSF against SARS-CoV-2 spike and receptor binding domain proteins using a multiplex microsphere assay. Internal validation showed that the saliva antibody assay had an estimated sensitivity of 76% and specificity of 100% based on 51 positive and 41 negative controls using oral fluid reference standards.

Test results from RT-PCR were available within two business days and were shared with participants via their preferred method of contact (phone, email, or text).

### Outcome measures

We used the proportion of eligible participants who enrolled in the study, and then subsequently completed self-collection at both time points, as a measure of acceptability. The research team qualitatively assessed testing feasibility in the camp setting through observations of proper test self-collection and camp staff willingness and capability to supervise testing.

The research team qualitatively observed adherence to written infection mitigation policies for each camp (Supplement D) regarding masking, cohort size, pod interactions, symptom screening, ventilation, and physical distancing on testing days.

RT-PCR and antibody testing results provide measures of acute incidence of infection and prevalence of prior infection, respectively, in the study population.

### Analysis

We report descriptive statistics of participant demographics, household characteristics, and RT-PCR and antibody test results, comparing proportions using chi-squared tests, means using t-tests, and medians using nonparametric equality-of-medians tests. We did not perform multivariable analyses of associations between participant characteristics and antibody results due to the low number of positive antibody tests.

We used Stata 16.1 (StataCorp LLC, College Station, TX). The institutional review board at UCSF approved this study as public health surveillance.

## Results

Of 246 eligible participants, 186 (76%) initially consented and enrolled. From the enrolled group, we excluded 6 campers and 17 household contacts from our analysis who were not present on both testing days **(Figure 1)**.

**Figure 1:**
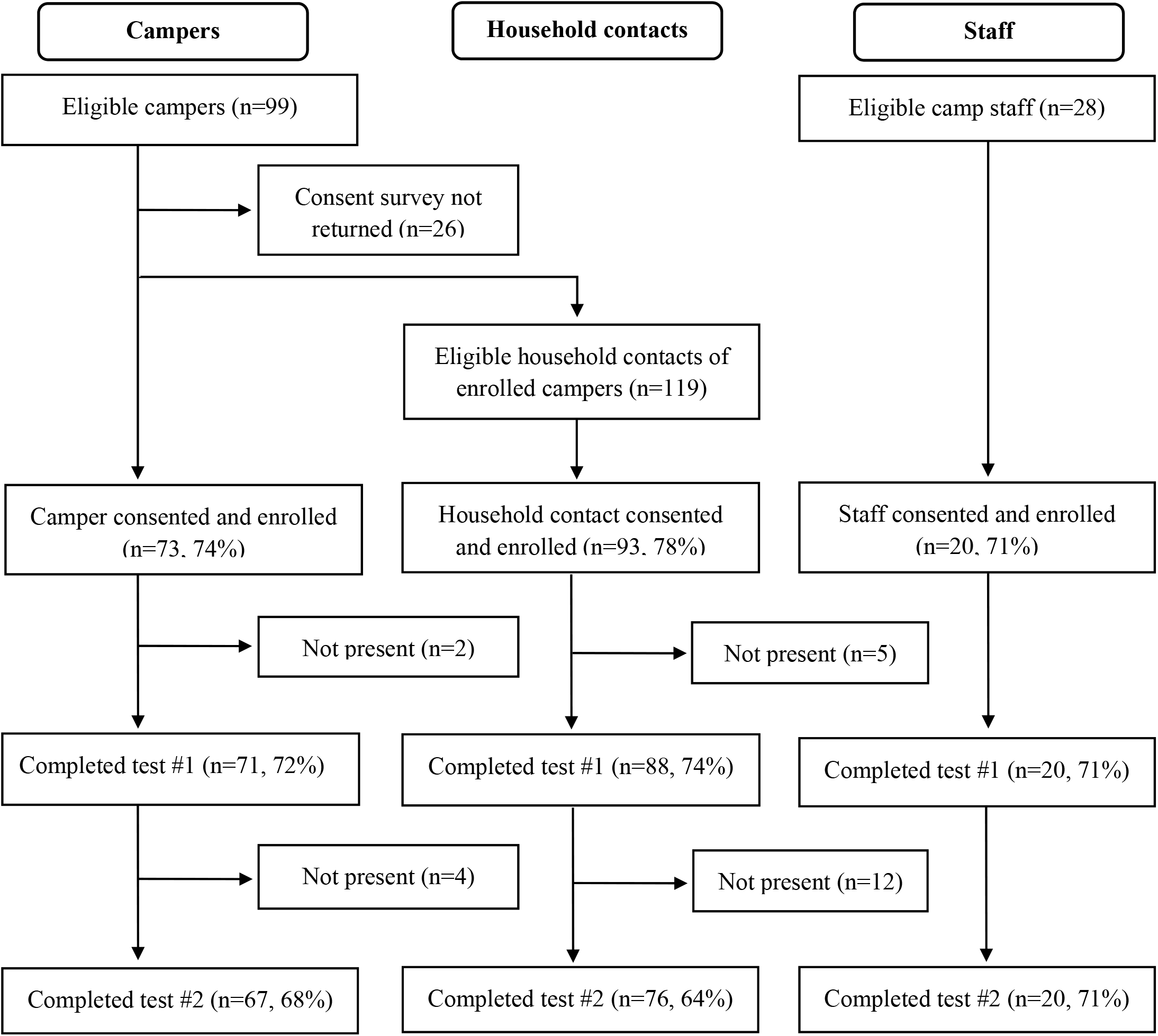
Flow Diagram of Eligible Population, Study Exclusions, and Final Cohort Selection

Our final cohort comprised 163 participants, including 67 campers, 76 household contacts, and 20 staff (**Table 1**). Campers in the final cohort were aged 5-14 (mean 9) years. More participants came from the science camp (55%, n=90). The sample was diverse, with white participants comprising less than half (48%, n=78). Preferred method of contact varied (phone call: 11% (n=6), email: 32% (n=18), text message: 57% (n=32)).

**Table 1:** Demographics for Participants Completing Both Rounds of Testing (N=163)

|  | **Campers** | **Household**<br>**Contacts** | **Staff** | **All** |
| --- | --- | --- | --- | --- |
| Participants, N | 67 | 76 | 20 | 163 |
| Camp, n (%) |  |  |  |  |
| General Camp | 30 (45) | 30 (40) | 13 (65) | 73 (45) |
| Science | 37 (55) | 46 (61) | 7 (35) | 90 (55) |
| Male, n (%)<sup>a</sup> | 34 (51) | 30 (40) | 7 (35) | 71 (44) |
| Age, mean (95% CI)<sup>b</sup> | 9 (9-10) | 46 (45-48) | 25 (20-29) | 28 (26-31) |
| Race/Ethnicity, n (%) |  |  |  |  |
| Caucasian | 27 (40) | 42 (55) | 9 (45) | 78 (48) |
| Latinx | 19 (28) | 23 (30) | 5 (25) | 47 (29) |
| Asian | 8 (12) | 8 (11) | 1 (5) | 17 (10) |
| Multiracial | 10 (15) | 0 (0) | 4 (20) | 14 (9) |
| Other, or decline | 3 (5) | 3 (4) | 1 (5) | 7 (4) |

CI, confidence interval
<sup>a</sup>One household contact was trans-male (included as male); one household contact did not respond and was not included;
<sup>b</sup>Age range was bi-modal among staff, with most staff 20-30 years of age, and several 40-50 years.

General camp households (n=25) were more likely to report at least one participant with Latinx ethnicity compared to science camp households (n=31) (76% vs. 3%, p<0.001; **Table 2**). General camp households were in areas with higher cumulative incidence of COVID-19 (mean 53 cases/10,000 people vs. 21 cases/10,000, p<0.001). There were no significant differences between the general day camp and science camp in median total household size (4 vs 5, p=0.80), nor in percentage of households with at least one frontline worker (32% vs 39%, p=0.61).

**Table 2:** Household Characteristics for Camp Participants & Household Contacts (N=56)

|  | All households | General Camp | Science Camp | p-value |
| --- | --- | --- | --- | --- |
| **Households, n (%)** | **56** | **25** | **31** | **--** |
| Family size, median (IQR) | 4.5 (4-5) | 4 (4-5) | 5 (4-6) | 0.80<sup>a</sup> |
| Latinx household, n (%)<sup>b</sup> | 20 (36) | 19 (76) | 1 (3) | <0.001<sup>c</sup> |
| COVID-19 incidence in household location, mean (95% CI)<sup>d</sup> | 35 (28 – 42) | 53 (43 – 62) | 21 (15 – 27) | <0.001<sup>e</sup> |
| Frontline worker household<sup>f</sup>, n (%) | 20 (36) | 8 (32) | 12 (39) | 0.61<sup>c</sup> |
| Healthcare | 6 (30) | 2 (25) | 4 (33) | 0.69<sup>c</sup> |
| Grocery or pharmacy | 0 (0) | 0 (0) | 0 (0) | -- |
| Construction | 4 (20) | 2 (25) | 2 (17) | 0.65<sup>c</sup> |
| Other | 12 (60) | 4 (50) | 8 (67) | 0.46<sup>c</sup> |
| Highest educational attainment in household, n (%) |  |  |  | 0.001<sup>c</sup> |
| Did not complete high school | 3 (5) | 3 (12) | 0 (0) | -- |
| High school | 4 (7) | 3 (12) | 1 (3) | -- |
| Some college | 7 (12) | 7 (28) | 0 (0) | -- |
| College | 13 (23) | 4 (16) | 9 (29) | -- |
| Graduate study | 26 (46) | 6 (24) | 20 (65) | -- |
| Decline/unsure | 3 (5) | 2 (8) | 1 (3) | -- |
| Adult diagnosed with COVID-19 in household before camp, n (%) | 1 (2) | 1 (4) | 0 (0) | 0.26<sup>c</sup> |
| Adult suspected to have COVID-19 in household before camp, n (%) | 3 (5) | 1 (4) | 2 (6) | 0.26<sup>c</sup> |

IQR, interquartile range; CI, confidence interval
<sup>a</sup>P-value calculated using nonparametric equality-of-medians test;
<sup>b</sup>Household with one or more Latinx participant compared to households without any Latinx participants;
<sup>c</sup>P-value calculated using chi-squared;
<sup>d</sup>Cumulative incidence of COVID-19 cases/10,000 people during June 19<sup>th</sup> - 21<sup>st</sup>, 2020 for household zip code or household city, if zip code data not available (n=8 households);
<sup>e</sup>P-value calculated using t-test;
<sup>f</sup>Percentages represent the percent of frontline workers within the category, and add up to over 100% if there are multiple frontline workers in same household. Across all households there were 22 frontline workers, eight from the general camp and 14 from the science camp.

### Acceptability and Feasibility of Specimen Collection

76% (186/246) of eligible participants consented to participate, including 74% (73/99) of campers; 71% (20/28) of staff, and 78% (93/119) of eligible adult household contacts (**Figure 1**). Of eligible participants, 163 (66%) successfully completed the serial testing approach and were included in the final cohort. Among those present, 100% of campers and staff completed testing at the second time point after participating in the first.

There was interest and availability for classroom staff to supervise campers in self-collection at the general day camp but this was not feasible at the science camp, due to staff remaining in classrooms with non-participating campers.

During supervised self-collection, younger children were most likely to need coaching, including in applying adequate force with the nasal swab, or not chewing and/or sucking on the saliva swab. During camp staff training, the most common corrections were assuring optimal camper self-collection and maintaining 6-foot distance from campers, with improvements noted over time.

### Infection mitigation policies and observations

Both camps had the following written policies: cohorts with ≤12 campers and 2 camp staff; separate classrooms and no interaction between pods; staff mask requirement at all times except while eating; and temperature checks on arrival (**Table 3**). The general day camp had additional policies: daily on-site symptom screening, 6 feet of physical distancing between campers in-cohort, and encouragement of camper masking. The science camp required camper masking at all times except for eating and had a policy of open windows and doors for ventilation (Supplement D).

**Table 3:** Summary of Camp Infection Mitigation Written Policies at Each Camp

| Policy | General Camp | Science Camp |
| --- | --- | --- |
| Staff Masking | **Required** | **Required** |
| Camper Masking | Encouraged | **Required** |
| Staff:camper cohort ratio | 2:12 | 2:12 |
| Temperature checks on site | **Required** | **Required** |
| Verbal symptom screening on site | **Required** | *Not required* |
| Physical Distancing in Cohort | **Required** | *Not required* |
| Ventilation and Open Windows | **Required** | **Required** |
| Hand Hygiene | Encouraged | Encouraged |
| High Touch Surface Disinfecting | **Required** | **Required** |

On observation, both camps’ cohort sizes were within stated goals. We observed consistent adherence with the science camp’s strict mask policy, and generally high but variable masking adherence at the general camp where masking was encouraged. There was variable consistency observed with cohorting policies at the science camp and consistent adherence at the general camp. There was variable adherence to the general camp’s within-cohort physical distancing policy, especially among younger children. Windows and doors were open in all classrooms in the science camp; ventilation observations were not conducted at general camp as testing occurred outdoors.

### Testing results

SARS-CoV-2 was not detected by RT-PCR at either time point among the 163 participants in the final cohort, nor was in detected in the tests of the 16 participants excluded for missing a testing time point. In all RT-PCR tests, the human RNAse P gene was detected, indicating adequate sample collection. RT-PCR follow-up results were unavailable for two participants whose specimen transport tubes from the second testing day leaked. All participants were asymptomatic at the time of SARS-CoV-2 testing, except one camper who was excluded from camp but who underwent self-testing outside the camp before returning home.

Within the final cohort, 7/163 participants (4%, 95% confidence interval 1-7%) had SARS-CoV-2 antibodies at one or more time point (four campers, two household contacts, and one staff, see **Table 4**, Supplements E and F). Of these, six (86%) were Latinx and from a high or moderate incidence zip code. There were two household clusters with detectable antibodies, each with one camper and one household contact. Three testing samples were lost, and one was indeterminate. One participant had antibodies detected at the end of camp but not the start, suggestive of seroconversion; this participant was general camp staff, living in a higher incidence zip code. Two participants reverted from positive to negative; both were children aged 7.

**Table 4:**
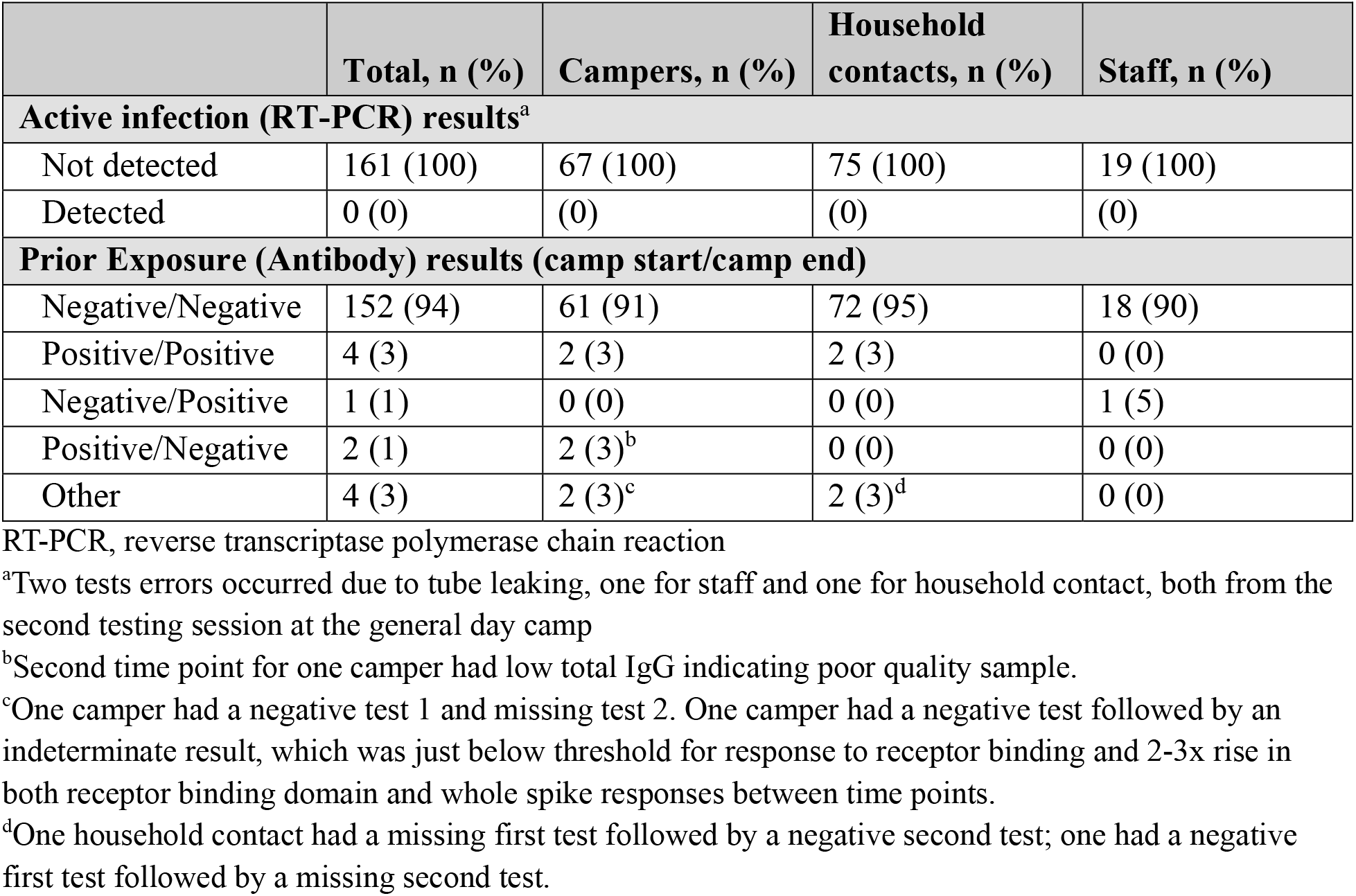
COVID-19 Active Infection and Prior Exposure Results, by Participant Type (N=163)

|  | **Total, n (%)** | **Campers, n (%)** | **Household contacts, n (%)** | **Staff, n (%)** |
| --- | --- | --- | --- | --- |
| **Active infection (RT-PCR) results<sup>a</sup>** |  |  |  |  |
| Not detected | 161 (100) | 67 (100) | 75 (100) | 19 (100) |
| Detected | 0 (0) | (0) | (0) | (0) |
| **Prior Exposure (Antibody) results (camp start/camp end)** |  |  |  |  |
| Negative/Negative | 152 (94) | 61 (91) | 72 (95) | 18 (90) |
| Positive/Positive | 4 (3) | 2 (3) | 2 (3) | 0 (0) |
| Negative/Positive | 1 (1) | 0 (0) | 0 (0) | 1 (5) |
| Positive/Negative | 2 (1) | 2 (3)<sup>b</sup> | 0 (0) | 0 (0) |
| Other | 4 (3) | 2 (3)<sup>c</sup> | 2 (3)<sup>d</sup> | 0 (0) |

RT-PCR, reverse transcriptase polymerase chain reaction
<sup>a</sup>Two tests errors occurred due to tube leaking, one for staff and one for household contact, both from the second testing session at the general day camp
<sup>b</sup>Second time point for one camper had low total IgG indicating poor quality sample.
<sup>c</sup>One camper had a negative test 1 and missing test 2. One camper had a negative test followed by an indeterminate result, which was just below threshold for response to receptor binding and 2-3x rise in both receptor binding domain and whole spike responses between time points.
<sup>d</sup>One household contact had a missing first test followed by a negative second test; one had a negative first test followed by a missing second test.

## Discussion

In this longitudinal study, we observed feasibility and acceptability of supervised serial self-collection for COVID-19 testing in urban, diverse, indoor camps in kindergarten through 8^th^ grade campers, their household contacts, and staff. Children as young as 5 years participated in self-collection at two time points, with 100% of consented and present campers and staff completing both rounds. Camp staff were able to assist with collection, demonstrating this as a possibility for schools, if needed. In addition, in the context of observed adherence to infection mitigation methods we found no evidence of active infections by RT-PCR but did find evidence of seroconversion in one young adult.

This study adds to existing evidence demonstrating promising acceptability and feasibility of less-invasive as well as self-collected SARS-CoV-2 testing strategies.^11,16^ Children age 6-18 years with cystic fibrosis were able to perform unsupervised anterior nares self-collection to test for common respiratory viruses; they reported high acceptability and the approach led to increased yield compared to provider-collected swabs.^16^ For SARS-CoV-2, multiple studies have demonstrated comparable results with self-collected testing compared to provider-collected NP sampling, including with supervised anterior nares self-collection,^9^ as well as unsupervised^10^ and supervised mid-turbinate self-collection.^17^ To our knowledge, this is the first study to describe self-collected anterior nares SARS-CoV-2 testing among children, or any participants in a school-like setting.

We noted potential limitations in the feasibility of self-collected saliva testing in younger children, including the need for closer coaching among younger children for adequate saliva sampling. The antibody reversion of two young children based on saliva samples may have been due to variable sample collection, as it is biologically unlikely that they would have reverted in this time frame, and one of the two had evidence of low total antibodies in the second sample. These findings emphasize the importance of feasibility assessments when introducing novel sampling strategies for children. For example, Wyllie et al. have demonstrated self-collected saliva RT-PCR testing SARS-CoV-2 testing to be feasible and to have excellent test characteristics in adults.^18^ However, this saliva sampling method requires collection prior to eating or drinking upon waking, and production of 3ml of saliva without bubbles into a cup, which is more complicated than the saliva collection we studied.

Neither indoor camp experienced documented COVID-19 cases or outbreaks in the context of described infection mitigation strategies, despite levels of community transmission greater than the California county “watchlist” criteria for school reopening (<10 new cases per 10,000 people per 14 days).^15^ This finding aligns with prior literature demonstrating limited or no school-based outbreaks or transmission in the setting of physical distancing, stable cohorts, reduced numbers of students, though prior studies had lower community incidence than in our study.^19-21^ The seroconversion of the single young adult in our study may indicate community acquisition. Other studies have described substantial outbreaks amongst those aged 6-19 years in the absence of adequate mitigation policies (specifically: large cohorts, poor ventilation, and no masking).^22,23^

Our findings have implications for potential school reopening and testing strategies. The methods we describe suggest a scalable approach for return to school testing. Current testing strategies utilizing provider-collected RT-PCR are limited by the need for skilled collectors, discomfort leading to increased likelihood of child and adult refusal, risk of transmission, and high utilization of PPE. We demonstrated a testing strategy that overcame these obstacles to permit serial test sampling in children as young as five years. While supervised self-collection is especially applicable to settings outside the clinic, these benefits also apply to healthcare environments.

From a public health standpoint, our results suggest the potential for school-based testing of asymptomatic individuals as part of school surveillance and contact tracing. The CDC notes that schools may perform school-based testing if they have capabilities to do so. Currently the CDC does not recommend surveillance of school-populations based on insufficient evidence weighed against the logistical barriers.^6^ An acceptable and feasible school-based testing strategy could potentially tip the scales toward this approach.

Finally, our study supports the feasibility of current infection mitigation strategies, including masking and stable cohorts in young children—interventions that will be key to limiting school-based transmission and enabling ongoing in-person education.^24-26^ The testing methodology we utilized could facilitate research to assess the efficacy of different infection mitigation strategies, with is sorely needed to inform policy on safe and successful education.

Our study has some limitations. Our setting was limited to two camps in San Francisco and camp duration was 3-5 weeks. The acceptability and feasibility we observed may not generalize to other areas of the nation, or to the longer duration of the school year. However, our relatively large sample of participants, including campers as young as 5 years, as well as consistent experience across camps with diverse populations support the potential generalizability. To date, clinical validation of self-testing has been limited to symptomatic adults. We did not compare the anterior nares self-collection in asymptomatic participants or children to the gold standard of healthcare personnel-collected NP swabs. It is possible that our approach had lower sensitivity than the gold standard and we may have missed some COVID-19 infections. However, given that viral levels appear to be highest before and during early symptom onset, and the nares have comparatively high viral loads, there is biologic plausibility for anterior nares collection in asymptomatic participants.^27,28^ In addition, the detection of human RNAse P gene in all samples suggests adequate sample collection and supports the validity of the negative test results. Of note, recent data suggest that frequent testing with short turnaround times, even with lower test sensitivity, is more likely to identify a subject early in COVID-19 infection, even while asymptomatic, than highly sensitive tests performed less frequently.^29^ Hence, the acceptability of the anterior nasal collection, allowing for frequent repeated testing, is potentially a key piece to preventing transmission.

In conclusion, we demonstrated excellent feasibility and acceptability of a serial surveillance SARS-CoV-2 testing approach with supervised anterior nares self-collection in 5-14 year-old children, their household contacts, and staff, during indoor summer camp. The test collection methods and infection mitigation strategies described here may be potential core components of safe school reopening.

## Data Availability

Data not currently posted for public viewing

## Acknowledgements

We thank the staff and leadership of the two summer camps for their close collaboration, with special acknowledgement to the leadership of Rochelle Celedon and Judith Diaz of the Buena Vista Day Camp. We thank Dr. Kelley Meade of Benioff Children’s Hospital Oakland for her contributions to the study design. We offer our gratitude to the families and camp staff for their participation. We thank the laboratory staff at the Chan Zuckerberg Biohub, collectively working as the CLIAhub consortium, who performed RT-PCR testing for our participants. The members of the CLIAhub consortium are: Irene Acosta, Vida Ahyong, Erika C. Anderson, Shaun Arevalo, Daniel Asarnow, Shannon Axelrod, Patrick Ayscue, Camillia S. Azimi, Caleigh M. Azumaya, Stefanie Bachl, Iris Bachmutsky, Aparna Bhaduri, Jeremy Bancroft Brown, Joshua Batson, Astrid Behnert, Ryan M. Boileau, Saumya R. Bollam, Alain R. Bonny, David Booth, Michael Jerico B. Borja, David Brown, Bryan Buie, Cassandra E. Burnett, Lauren E. Byrnes, Katelyn A. Cabral,, Joana P. Cabrera, Saharai Caldera,, Gabriela Canales, Gloria R. Castañeda, Agnes Protacio Chan, Christopher R. Chang, Arthur Charles-Orszag,, Carly Cheung, Unseng Chio, Charles Chiu, Eric D. Chow, Y. Rose Citron, Allison Cohen, Lillian B. Cohn,, Mitchel A. Cole, Daniel N. Conrad, Angela Constantino, Andrew Cote, Tre’Jon Crayton-Hall, Spyros Darmanis, Angela M. Detweiler, Rebekah L. Dial, Shen Dong, Elias M. Duarte, David Dynerman, Rebecca Egger, Alison Fanton, Stacey M. Frumm, Becky Xu Hua Fu, Valentina E. Garcia, Julie Garcia, Christina Gladkova,, Miriam Goldman, Rafael Gomez-Sjoberg, M. Grace Gordon, James C.R. Grove, Shweta Gupta, Alexis Haddjeri-Hopkins, Pierce Hadley,, John Haliburton, Samantha L. Hao, George Hartoularos, Nadia Herrera, Melissa Hilberg, Kit Ying E. Ho, Nicholas Hoppe, Shayan Hosseinzadeh, Conor J. Howard, Jeffrey A. Hussmann, Elizabeth Hwang, Danielle Ingebrigtsen, Julia R. Jackson, Ziad M. Jowhar, Danielle Kain, James Y.S. Kim, Amy Kistler, Oriana Kreutzfeld, Jessie Kulsuptrakul, Andrew F. Kung, Charles Langelier,, Matthew T. Laurie, Lena Lee, Kun Leng, Kristoffer E Leon, Manuel D. Leonetti, Sophia R. Levan, Sam Li, Aileen W. Li, Jamin Liu, Heidi S. Lubin, Amy Lyden, Jennifer Mann, Sabrina Mann, Gorica Margulis, Diana M. Marquez, Bryan P. Marsh, Calla Martyn, Elizabeth E. McCarthy, Aaron McGeever, Alexander F. Merriman, Lauren K. Meyer, Steve Miller, Megan K. Moore, Cody T. Mowery, Tanzila Mukhtar, Lusajo L. Mwakibete, Noelle Narez, Norma F. Neff, Lindsay A. Osso, Diter Oviedo, Suping Peng, Maira Phelps, Kiet Phong, Peter Picard, Lindsey M. Pieper, Neha Pincha, Angela Oliveira Pisco, Angela Pogson, Sergei Pourmal, Robert R. Puccinelli, Andreas S. Puschnik, Elze Rackaityte, Preethi Raghavan, Madhura Raghavan, James Reese, Joseph M. Replogle, Hanna Retallack, Helen Reyes, Donald Rose, Marci F. Rosenberg, Estella Sanchez-Guerrero, Sydney M. Sattler, Laura Savy, Stephanie K. See, Kristin K. Sellers, Paula Hayakawa Serpa,, Maureen Sheehy, Jonathan Sheu, Sukrit Silas, Jessica A. Streithorst, Jack Strickland, Doug Stryke, Sara Sunshine, Peter Suslow, Renaldo Sutanto, Serena Tamura, Michelle Tan, Jiongyi Tan, Alice Tang, Cristina M. Tato, Jack C. Taylor, Iliana Tenvooren, Erin M. Thompson, Edward C. Thornborrow, Eric Tse, Tony Tung, Marc L. Turner, Victoria S. Turner, Rigney E. Turnham, Mary J. Turocy, Trisha V. Vaidyanathan, Ilia D. Vainchtein, Manu Vanaerschot, Sara E. Vazquez, Anica M. Wandler, Anne Wapniarski, James T. Webber, Zara Y. Weinberg, Alexandra Westbrook, Allison W. Wong, Emily Wong, Gajus Worthington, Fang Xie, Albert Xu, Terrina Yamamoto, Ying Yang, Fauna Yarza, Yefim Zaltsman, Tina Zheng

## Abbreviations

CDC: Centers for Disease Control and Prevention;
COVID-19: coronavirus disease 2019;
NP: nasopharyngeal;
PPE: personal protective equipment;
RT-PCR: reverse transcription polymerase chain reaction;
SARS CoV-2: severe acute respiratory syndrome coronavirus 2;
UCSF: University of California, San Francisco.

## References

1. Levinson M, Cevik M, Lipsitch M. Reopening primary schools during the pandemic. N Engl J Med. 2020:684–688. doi:10.1056/NEJMms2024920

2. Donohue JM, Miller E. COVID-19 and school closures. JAMA. 2020:1–3. doi:10.1001/jama.2020.13092

3. Schleicher A. The Impact of Covid-19 on education insights From Education At a Glance 2020. http://www.oecd.org/education/the-impact-of-covid-19-on-education-insights-education-at-a-glance-2020.pdf?utm_source=COVID-19+Policy+Update&utm_campaign=1653126165-EMAIL_CAMPAIGN_2020_09_09_02_47&utm_medium=email&utm_term=0_b6a8e3fa3e-1653126165-423107933. Published 2020. Accessed September 18, 2020.

4. Fuchs-Schündeln N, Krueger D, Ludwig A, Popova I. The Long-Term Distributional and Welfare Effects Of COVID-19 School Closures. Vol 47. Cambridge, MA; 2020. doi:10.1017/CBO9781107415324.004

5. Dorn E, Hancock B, Sarakatsannis J, Viruleg E. COVID-19 and student learning in the United States: The hurt could last a lifetime. McKinsey & Company. https://www.mckinsey.com/industries/public-and-social-sector/our-insights/covid-19-and-student-learning-in-the-united-states-the-hurt-could-last-a-lifetime?referringSource=articleShare#. Published 2020. Accessed September 18, 2020.

6. Centers for Disease Control and Prevention. Interim considerations for K-12 school administrators for SARS-CoV-2 testing. https://www.cdc.gov/coronavirus/2019-ncov/community/schools-childcare/k-12-testing.html. Published 2020. Accessed August 18, 2020.

7. Fitzsimmons EG. N.Y.C. school plan hinges on hundreds of thousands of virus tests. New York Times. https://www.nytimes.com/2020/09/02/nyregion/schools-reopen-testing-nyc.html. Published September 2, 2020.

8. United Federation of Teachers. Testing & tracing. https://www.uft.org/your-rights/safety-health/coronavirus/faq-on-school-reopening/testing-tracing. Published 2020. Accessed September 11, 2020.

9. Tu Y-P, Jennings R, Hart B, et al. Swabs collected by patients or health care workers for SARS-CoV-2 testing. N Engl J Med. 2020:1–3. doi:10.1056/nejmc2016321

10. McCulloch DJ, Kim AE, Wilcox NC, et al. Comparison of unsupervised home self-collected midnasal swabs with clinician-collected nasopharyngeal swabs for detection of SARS-CoV-2 infection. JAMA Netw Open. 2020;3(7):1–4. doi:10.1001/jamanetworkopen.2020.16382

11. Frazee BW, Rodríguez-Hoces de la Guardia A, Alter H, et al. Accuracy and discomfort of different types of intranasal specimen collection methods for molecular influenza testing in emergency department patients. Ann Emerg Med. 2018;71(4):509-517.e1. doi:10.1016/j.annemergmed.2017.09.010

12. Centers for Disease Control and Prevention. Overview of testing for SARS-CoV-2. http://web.archive.org/web/20200818231652/https://www.cdc.gov/coronavirus/2019-ncov/hcp/testing-overview.html. Published 2020. Accessed August 18, 2020.

13. Centers for Disease Control and Prevention. Interim guidelines for collecting, handling, and testing clinical specimens from persons for Coronavirus Disease 2019 (COVID-19). https://web.archive.org/web/20200817202748/https://www.cdc.gov/coronavirus/2019-ncov/lab/guidelines-clinical-specimens.html. Accessed August 18, 2020.

14. Emmons WW, Paparello SF, Decker CF, Sheffield JM, Lowe-Bey FH. A modified elisa and western blot accurately determine anti-human immunodeficiency virus type 1 antibodies in oral fluids obtained with a special collecting device. J Infect Dis. 1995;171(6):1406–1410. doi:10.1093/infdis/171.6.1406

15. Los Angeles Times. California Coronavirus Data. http://web.archive.org/web/20200821002734/https://github.com/datadesk/california-coronavirus-data/blob/master/latimes-place-totals.csv. Published 2020. Accessed August 20, 2020.

16. Emerson J, Cochrane E, McNamara S, Kuypers J, Gibson RL, Campbell AP. Home self-collection of nasal swabs for diagnosis of acute respiratory virus infections in children with cystic fibrosis. J Pediatric Infect Dis Soc. 2013;2(4):345–351. doi:10.1093/jpids/pit039

17. Wehrhahn MC, Robson J, Brown S, et al. Self-collection: An appropriate alternative during the SARS-CoV-2 pandemic. J Clin Virol. 2020;128(April):104417. doi:10.1016/j.jcv.2020.104417

18. Wyllie AL, Fournier J, Campbell ACM, Tokuyama M, Vijayakumar P. Saliva or nasopharyngeal swab specimens for detection of SARS-CoV-2. N Engl J Med. 2020:1–4. doi:10.1056/NEJMc2016359

19. Carlson J. Covid-19 in schoolchildren: a comparison between Finland and Sweden. https://www.folkhalsomyndigheten.se/contentassets/c1b78bffbfde4a7899eb0d8ffdb57b09/covid-19-school-aged-children.pdf. Published 2020. Accessed August 3, 2020.

20. National Centre for Immunisation Research and Surveillance (NCIRS). COVID-19 in schools – the experience in NSW. 2020;9(April):1–5.

21. Link-Gelles R, DellaGrotta AL, Molina C, et al. Limited secondary transmission of SARS-CoV-2 in child care programs — Rhode Island, June 1–July 31, 2020. MMWR Recomm Rep. 2020;69(34):1170–1172. doi:10.15585/mmwr.mm6934e2

22. Szablewski CM, Chang KT, Brown MM, et al. SARS-CoV-2 transmission and infection among attendees of an overnight camp-Georgia, June 2020. MMWR Recomm Rep. 2020;69(31):2019–2021. doi:10.15585/mmwr

23. Stein-Zamir C, Abramson N, Shoob H, et al. A large COVID-19 outbreak in a high school 10 days after schools’ reopening, Israel, May 2020. Euro Surveill. 2020;25(29):1– 5. doi:10.2807/1560-7917.ES.2020.25.29.2001352

24. Gandhi M, Beyrer C, Goosby E. Masks do more than protect others during COVID-19: reducing the inoculum of SARS-CoV-2 to protect the wearer. J Gen Intern Med. 2020:17–20. doi:10.1007/s11606-020-06067-8

25. Gandhi M, Rutherford GW. Facial masking for COVID-19 — potential for “variolation” as we await a vaccine. N Engl J Med. 2020:1969–1973. doi:DOI: 10.1056/NEJMp2009027

26. Head JR, Andrejko K, Cheng Q, et al. The effect of school closures and reopening strategies on COVID-19 infection dynamics in the San Francisco Bay Area: a cross-sectional survey and modeling analysis. medRxiv. 2020;1(510):2020.08.06.20169797. doi:10.1101/2020.08.06.20169797

27. Zou L, Ruan F, Huang M, et al. SARS-CoV-2 viral load in upper respiratory specimens of infected patients. N Engl J Med. 2020;382(12):1175–1177. doi:10.1056/NEJMc2000231

28. He X, Lau EHY, Wu P, et al. Temporal dynamics in viral shedding and transmissibility of COVID-19. Nat Med. 2020;26(5):672–675. doi:10.1038/s41591-020-0869-5

29. Larremore DB, Wilder B, Lester E, et al. Test sensitivity is secondary to frequency and turnaround time for COVID-19 surveillance. medRxiv. 2020:2020.06.22.20136309. doi:10.1101/2020.06.22.20136309

