## Supplemental Materials for "Supervised self-collected SARS-CoV-2 testing in indoor summer camps to inform school reopening"

**Supplements**:

Supplement A: Instructional video of anterior nares self-collection

Supplement B: Photo of camper performing supervised self-collection

Supplement C: Standard operating procedures for swab collections

Supplement D: Written infection control protocols at two camps, distributed to parents and staff

Supplement E:  Table of associations between antibody results and participant demographics

Supplement F:  Description of participants with any antibody results, categorized by status at beginning and end of camp status (Positive/Positive, Negative/Positive, Positive/Negative)

**Supplement A:** Instructional video of anterior nares self-collection

**Video 1:** Instructional video of anterior nares self-collection

**Supplement B:** Camper performing supervised self-collection

**Figure 2:** Camper performing supervised self-collection

**Supplement C:** Standard operating procedures for swab collections

Participants performed hand hygiene before and after specimen collection using alcohol-based hand sanitizer. Specimen collectors performed hand hygiene and changed gloves for each participant. Specimen collectors opened swab packages and presented the swab to the participant in the opened package, without handling the swab. Then specimen collectors stepped back to allow 6 feet of distance between themselves and the participant, instructing the participant to remove their mask. For anterior nares self-collection, participants inserted the flocked swab about 0.5 inch deep and rubbed around the inner walls of both nostrils for 10 seconds each (Figure 2, instructional video linked in Supplement A).[13] Participants then handed the swab back to the specimen collector, who placed the swab into DNA/RNA Shield (Zymo research, Irvine, CA) for stabilization during transport. Saliva was collected using the OraSure Oral Specimen Collection Device (OraSure Technologies, Bethlehem, PA). Participants took the swab and rubbed and then retained the swab against their outer gum line for at least two minutes, timed by the specimen collector. The gum line has higher concentration of antibodies with fewer inhibitory substances compared to other parts of the mouth.[14] Participants then handed the swab back to the specimen collector who placed it in a separate OraSure collection tube.

**Supplement D:** Written infection control protocols at two camps, distributed to parents and staff

**General Day Camp**

Drop Off Procedures:

- Strict Drop Off Time: 8:30 a.m. - 8:45 a.m. Arrival time staggered for families. Please arrive promptly.
- Stand in designated marked spot on sidewalk with your child to maintain 6 feet distance
- All children will be screened with health questions and temperature taken
- Sign your child in with your own pen if possible
- Wash your hands and your child's hands before coming to program
- Do not come into the building. Arrive with a mask on, wait at the entry for a staff member to let your child in

Pick Up Procedures:

- Children must be picked up on time, between 3:00 p.m - 3:15 p.m
- Sign your child out with your own pen if possible
- Do not enter the school building. A staff member will be outside during dismissal time
- The staff member will call for your child to be escorted to you

Early Pick Up:

- Notify staff at drop-off if you must have an early pick-up. Staff will coordinate with parents' arrival and walk the child outside to them.

Classroom (pods):

- Maximum capacity is 12 children and 2 teachers, the same children and teachers everyday, every week
- Pods (classrooms) will not interact with other pods
- Each child will have a designated desk area that is 6 feet away from other children
- Each child is encouraged to wear a mask
- Hand washing/sanitizer will happen often during the day
- Classrooms will be cleaned often when children are not present in the classroom
- Windows opened and fans used for ventilation

Lunch:

- Children will eat lunch at staggered times

- Handwashing before and after eating lunch
- Absolutely no sharing food

Outdoor Play:

- Games that do not require contact will be played
- All equipment will be cleaned after each use

Bathroom Use:

- One child at a time will be escorted by an adult to the bathroom, adult will wait outside for the child
- Bathrooms will be cleaned after each use by an adult

Illness:

- Children with 100.4°F will be sent home and can return to Summer Camp after not having a fever for 72 hours.
- If a child has COVID-19 or has a household member who has COVID-19 they will not be able to return to camp for 14 days or when the Department of Health has given the OK.

What to do if your child is sick:

- If a child tests positive for COVID-19 we will shut down for 3-5 days to do a thorough cleaning of the camp area.
- The child may not return to camp for 14 days.
- If a child has a common cold with a fever they may not return to camp until they have not had a fever for 72 hours.
- If a child becomes sick during the day they will be kept in a separate but supervised space until they can go home. We will limit the number of staff who take care of sick children.

For staff:

- All staff will be screened with health questions and temperature at arrival.
- If you are sick please stay home. If you have a fever of 100.4 degrees you can not return to work until you have not had a fever for 72 hours
- If you or a member of your household has tested positive for COVID-19 please notify one of the administrators immediately. You will not be able to return to work for a 14 day period

Science Camp:

Classrooms:

- Campers will be grouped in “pods” according to their session.
- Campers will stay with their same pod for the entire three-week session.
- Pod size will be limited to 10 campers with some up to 12, depending on the size of the room
- Each pod will remain in a separate room and will not interact with kids from other pods.
- Two instructors will lead each pod and will remain with the same pod each week.
- Children and youth must attend the first week of the session. Those who do not attend the first week may not join the camp later and will not receive any refund or credit (SFDPH’s rule)
- Children will wear a facemask in classroom at all times. We will provide one washable three-layer soft cotton mask for each child for free. They are very breathable. Masks are washed every Wednesday and Friday. Masks will bear the name of each child and will be kept in an individual mask holder in the classroom for the duration of the session.
- Campers will take home their mask at the end of the session. We will use encouragement and appeal to reason when asking kids to wear masks.
- If a child manages to lose their mask somehow, a new one will be issued, and your account will be charged $5.
- If you would like your child to use his/her own mask, it needs to be reusable and have at least 2 layers of fabric.
- Children will wash hands for 20 seconds between activities, before eating, after recess, and before leaving for home; approximately 7 times a day.
- Children will sanitize surfaces and objects in the classroom twice a day. Including doorknobs, light switches, classroom sink handles, countertops, desks, chairs, and objects such as PCR machines, keyboards, and polishing machines.
- Staff will sanitize surfaces in the common areas twice daily.
- Classroom windows and doors will remain open as much as possible.

<u>Sign-in rules</u>

- We ask that you consider dropping off children in grades 2-5 at 8:45-9am, and grades 6-8 at 9-9:15. We know this is not always possible, but please try. This will help minimize movement and interaction.
- Family members and caregivers waiting outside to drop-off or pick-up children must wear face masks per SFDPH’s rules.
- We are sorry, but parents are not allowed to enter the building. If you have a shy small child, you may enter the lobby area only, wearing a face mask.
- Staff should remain 6 feet apart from parents and caregivers. Please show our staff you care and move about with this in mind.
- A staff member will take children’s temperatures with a thermometer upon arrival, using a “non-touch” (infrared) thermometer

- Sign in and out will be done using an app—detailed information to be sent separately.
- During AM drop off, a hygiene station will be located near the entrance for children and staff to use immediately upon their arrival.
- Please try not to have grandparents pick up campers. If you love someone over the age of 60, keep them away from busy places.
- Children with symptoms or fever will be sent home. Information about getting tested can be found here: https://sf.gov/find-out-how-get-tested-coronavirus

Children – Recess

- All lunch and snacks should be provided by the parents. [The Camp] will not be providing any snacks or lunch this year per regulations. Please make sure you pack enough food for your child.
- Children should not share food. Lunch and snacks will be taken in either the classroom. There will be no cafeteria this year, regrettably.
- Sports with shared equipment or physical contacts, like soccer and baseball, will be played only within the same pod.
- Campers in each pod will wear over their shirts, a Dry-FIT T-shirt with a unique color of their group. Shirts will be laundered by the camp at the end of each week and will be taken home at the end of the session.

**Supplemental Table E:** Associations between Antibody Results and Participant Demographics

| **Antibody test result** | **Negative (no positive Ab test)** | **Positive (any positive Ab test)** |
|---|---|---|
| All Participants | 156 | 7 |
| Participant Type, n (%) | | |
| Campers | 63 (40) | 4 (57) |
| Household contacts | 74 (47) | 2 (29) |
| Staff | 19 (12) | 1 (14) |
| Camp type, n (%) | | |
| General Day | 67 (43) | 6 (86) |
| Science | 89 (57) | 1 (14) |
| Gender, n (%)[a] | | |
| Male | 69 (44) | 2 (29) |
| Female | 85 (55) | 5 (71) |
| Age, years | | |
| 0-14 | 63 (40) | 4 (57) |
| 15-29 | 18 (12) | 1 (14) |
| 30+ | 75 (48) | 2 (29) |
| Race/Ethnicity, n (%) | | |

| | | |
|---|---|---|
| Not Latinx | 107 (69) | 1 (14) |
| Latinx | 49 (31) | 6 (86) |
| COVID incidence in household locality, mean (95% CI)[b] | 36 (32 – 40) | 53 (34 – 72) |
| Frontline worker in household, n (%) | 52 (33) | 3 (43) |
| Household member with history of suspected or confirmed COVID-19, n (%) | 8 (5) | 2 (29) |
| Household member antibody positive, n (%) | 1 (1.28) | 4 (57.1) |

Ab, antibody; CI Confidence Interval
[a]One household contact was trans male; one household contact did not respond
[b]Cumulative incidence of COVID-19 cases/10,000 people during June 19th - 21st, 2020 for household zip code (or city, for n=8 households).

**Supplement F:** Description of Participants with Any Antibody Results, Categorized by status at beginning and end of camp status (Positive/Positive, Negative/Positive, Positive/Negative)

Of the seven antibody positive participants, four had positive tests at both time points (sero-positive), one had an initial negative test at camp beginning and subsequent positive at camp end (sero-conversion), two had an initial positive test and a subsequent negative (sero-reversion). *Sero-positive*: The four with both tests positive comprised two clusters, each with a camper and adult contact in the same household. Both were in Latinx households in high or moderate incidence zip codes (71/10,000 and 47/10,000). One camper was under 10 years old, the other was older than 10 years. One cluster lived with a construction worker in the household. *Sero-conversion*: The participant who sero-converted was a Latinx staff member also in a high incidence zip code (71/10,000). *Sero-reversion*: The participants who reverted from positive to negative were both campers under 10 years old. We hypothesize they may not have given good samples in the second test day, as it is unlikely that they would have reverted in this time frame. No household contacts were tested for those who reverted. Both lived in households without any frontline workers, one in a low incidence zip code (13/10,000) and one in a moderate incidence zip code (50/10,000).
